# Forecasting Transmission Dynamics of COVID-19 Epidemic in India Under Various Containment Measures- A Time-Dependent State-Space SIR Approach

**DOI:** 10.1101/2020.05.08.20095877

**Authors:** Vishal Deo, Anuradha R. Chetiya, Barnali Deka, Gurprit Grover

## Abstract

**Objectives:** Our primary objective is to predict the dynamics of COVID-19 epidemic in India while adjusting for the effects of various progressively implemented containment measures. Apart from forecasting the major turning points and parameters associated with the epidemic, we intend to provide an epidemiological assessment of the impact of these containment measures in India.

**Methods:** We propose a method based on time-series SIR model to estimate time-dependent modifiers for transmission rate of the infection. These modifiers are used in state-space SIR model to estimate reproduction number *R_0_*, expected total incidence, and to forecast the daily prevalence till the end of the epidemic. We consider four different scenarios, two based on current developments and two based on hypothetical situations for the purpose of comparison.

**Results:** Assuming gradual relaxation in lockdown post 17 May 2020, we expect the prevalence of infecteds to cross 9 million, with at least 1 million severe cases, around the end of October 2020. For the same case, estimates of *R_0_* for the phases no-intervention, partial-lockdown and lockdown are 4.46 (7.1), 1.47 (2.33), and 0.817 (1.29) respectively, assuming 14-day (24-day) infectious period.

**Conclusions:** Estimated modifiers give consistent estimates of unadjusted *R_0_* across different scenarios, demonstrating precision. Results corroborate the effectiveness of lockdown measures in substantially reducing *R_0_*. Also, predictions are highly sensitive towards estimate of infectious period.

## 1 Introduction

### 1.1 Context

In the absence of vaccines or effective antiviral therapies for COVID-19, governments all over the world are turning to classical non-pharmaceutical public health measures to contain the epidemic, such as isolation, quarantine, social distancing and community containment. Rigorous implementation of these four traditional counter-measures helped in halting the earlier epidemic of SARS-CoV in 2002-2003 [Kundapur *et al*. (2020); Wilder-Smith and Freedman, (2020)]. As of May 8, 2020, infections across India have surged past 53,000 cases with 1783 deaths reported. The government of India has implemented these containment and mitigating interventions along with travel restrictions and lockdown of the entire country to slow down the spread of the virus. Epidemiological assessment using infectious disease modelling is the key to evaluate the impact of these measures on transmission dynamics of COVID-19 in India, and thus provide crucial information to the policy makers in government organizations to plan ahead for an effective and sustained public health response to manage the epidemic.

### 1.2 Review of epidemiological modeling of COVID-19

Recent studies of COVID-19 have attempted to predict number of case counts, rate of transmission, reproduction rate/ number (*R_0_*), size of epidemic and end date of the epidemic. *R_0_* is an important factor for risk assessment of any epidemic and is defined as the expected number of secondary cases that arise from a typical infectious index-case in a completely susceptible host population. Wang *et al*. (2020) have developed a health informatics toolbox with an R package called eSIR to understand epidemiological trend of COVID-19 in Hubei province and other regions of China. Their model considers a time varying quarantine factor to forecast future trend of COVID-19 spread in these regions. An earlier study by Chinazzi *et al*. (2020) assessed the impact of such restrictions based on data of over 3200 sub-populations in roughly 200 different countries and territories across the world. They have used a meta-population network approach, in which each sub-population is modelled using a Susceptible-Latent-Infectious-Removed (SLIR) model.

#### COVID-19 in India

Using a compartmental SEIR model, Chatterjee *et al*. (2020a) have concluded that effective implementation of quarantine and other non-pharmacological interventions would bring down the epidemic spread of COVID-19 in India to a manageable level. Mandal *et al*. (2020) used a SEIR model with a quarantine component to predict an effective reduction in cumulative incidence in India. A SIR model is developed by Singh and Adhikari (2020) based on data up to the first phase of India’s total lockdown to illustrate the need of sustained lockdowns with periodic relaxations. Some other recent work on COVID-19 in India include Tiwari (2020) and Gupta *et al*. (2020).

All these studies on COVID-19 infection dynamics in India are based on the assumption of constant disease transmission rate. However, phase-wise imposition of travel restrictions, lockdown and other non-pharmaceutical preventive measures, as well as increasing community level awareness with time, are expected to induce time varying effects in the transmission rate.

### 1.3 Our approach

To account for variations in transmission rate of the infection due to the implementation of various containment protocols, we propose to implement a time-dependent state-space SIR model to the observed data from India. Instead of taking a pre-specified step function modifier like Ray *et al*. (2020), we propose a time-series SIR based approach to estimate the phase-wise transmission modifiers. Modifier functions, both step and exponential, are estimated using the daily prevalence data reported in India from 2^nd^ March, 2020 to 30^th^ April, 2020.

## 2 Methodology

### 2.1 The extended state-space SIR model with time varying transmission rate

Several containment measures have been implemented in India at different points in time creating phases of quarantine/ containment levels across the country. Such phases are expected to exhibit different rates of transmission of the disease, *i.e*., transmission rates become time-dependent. If we assume that the change (or reduction) in the transmission rate is strictly because of macro level measures implemented by the authorities, we can define a specific transmission rate for each phase based on the level of containment. Wang *et al*. (2020) have proposed a step function approach to define such modifiers. However, it is also true that apart from the containment measures implemented by the government, rising awareness at micro community levels also contributes towards reducing the rate of transmission. To incorporate this idea, they have suggested defining the transmission modifier as a continuous function of time.

Suppose there are three different phases, with two points of major changes in quarantine/lockdown protocols. Let *P_i_* denotes i-th phase, such that *P_1_* represents the initial phase without any such protocol in place. Then, the step function for transmission rate modifier, *π(t)*, can be expressed as follows.

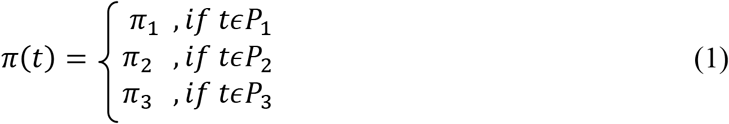

Where, *π_1_* = 1 if *P_1_* represents the phase without any intervention. Following exponential modifier functions can be used to account for continuous changes in modifier values with time.

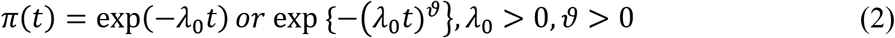

We have applied both approaches to define modifiers for the base transmission rate *β*. The effective rate of transmission at time t is given as, *β_t_ = β.π(t)*. Using this time-dependent transmission rate, the eSIR model proposed by Wang *et al*. (2020) is fitted to predict daily prevalence of infected, removed and susceptible. This model is a time-dependent version of the state-space SIR model introduced by Osthus *et al*. (2017), and can be defined as follows.

#### Model description

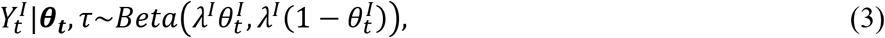

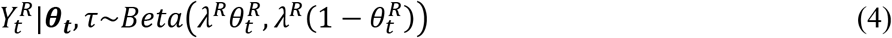

Where:

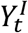 - Time series of proportion of infected cases

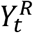 - Time series of proportion of removed cases (Recovered + Death)

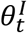 - Prevalence of infection at time t in terms of probability (probability of a person being infected at time t)

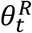 - Prevalence of removal at time t (probability of a person being removed from the infected compartment)

Also, the constants λ*^I^* and λ*^R^* control the variances of the respective observed proportions.

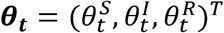 represents the latent population prevalence. It is a three-state Markov process where 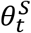 is the probability of a person being susceptible at time t. The Markov process (or the distribution of the transmissions of the Markov process) is defined as follows,

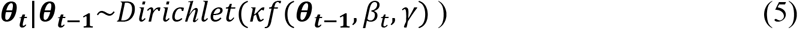

Thus the complete model is a Dirichlet-Beta state-space model. Here, *β_t_ = β.π*(*t*) is the modified/ adjusted contact rate multiplied by the probability of transmission given a contact between a susceptible and an infectious individual. The function f (.) in the argument of Dirichlet function is the SIR model given as follows.

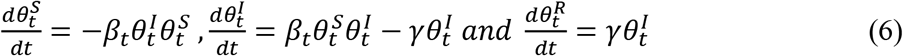

Solution of this set of differential equations is achieved using the Runge-Kutta approximation.

Overall success of this modelling structure depends heavily on the relevance of the modifier values specified for different phases. Using appropriate values of *π_i_’s* in (1) and the constants λ_0_ and *ϑ* in (2) will be imperative towards achieving reliable predictions. To avoid misleading predictions resulting from speculative pre-specified values of the modifiers, we propose methods based on time-series SIR model to estimate these values. The proposed method is described in the following section.

### 2.2 Estimation of modifiers of β for different phases of quarantine/ lockdown measures

Time-series SIR (TSIR) model [Bjørnstad *et al*. (2002); Finkenstadt, *et al*. (2002); Grenfell *et al*. (2002)] is used to estimate time-dependent modifier values. In the step function π(t), the steps (or phases) are defined according to different levels of preventive measures implemented by the government over the observed period of time. In TSIR model, the response, being a count variable, is assumed to follow certain discrete count process distribution like Poisson distribution or Negative Binomial distribution; refer Bjørnstad (2018). The basic structure of TSIR model can be defined as follows.

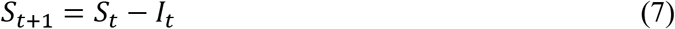

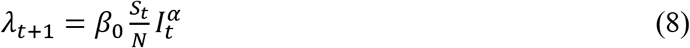

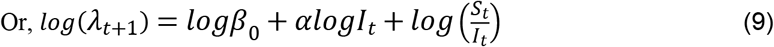

Where, *S_t_* and *I_t_* are the number of susceptibles and infecteds at time *t, N* is the population size, *β_0_* is the transmission rate and *λ_t+_*_1_ is the expected number of new infecteds at time *t*+1. New number of infecteds is assumed to follow Negative Binomial (or Poisson) distribution and a generalized Negative Binomial (or Poisson) linear model with log link is fitted with *logI_t_* as a covariate and 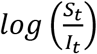 as an offset variable. The exponent *α* is expected to be just under 1 (*i.e*. close to 1) and is meant to account for discretizing the underlying continuous process. However, we can present an alternative interpretation of *α*, using basic SIR model, by writing an approximate expression (taking *α* = 1) for expected number of new infecteds at time *t+*1 with a time-varying transmission rate, as given below.

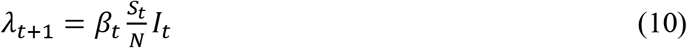

Comparing equations (8) and (10), we can see that if *α* = 1 (or close to 1), *β_t_* = *β_0_* (constant over time). However, if the value of *α* deviates considerably from 1, it has impact on the effective value of transmission rate, thus making the effective rate of transmission time-dependent. That is, in such cases *α* assimilates the changes in transmission rate over time. From equations (8) and (10), we can further write,

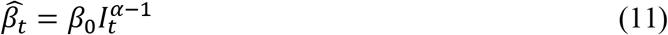

#### Option 1: Defining step function for phase specific modifiers

Using the TSIR model, we estimate *β_0_* and *α* separately for each phase. The effective transmission rate, *β_t_*, is then estimated at each time *t* using equation (11). Average of these estimates over the time range of a phase is taken as an estimate of the effective transmission rate for that phase. Suppose we have three time phases in our study, say *P_1_, P_2_*, and *P_3_*. Then, the estimate of phase specific transmission rate will be given as,

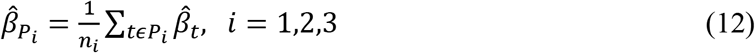

And the estimated step function of modifiers will be,

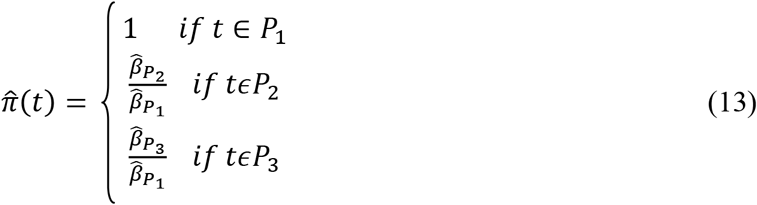

#### Option 2: Defining continuous time-dependent exponential modifier function

Instead of fitting phase-wise models, we fit a generalized linear model on the entire observed data and obtain estimates of effective transmission rates, 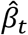, using equation (11). We derive estimates of modifiers at each time point *t* for the entire observed period as,

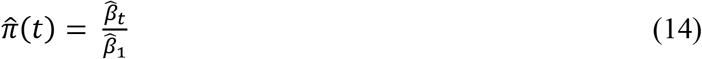

Where, 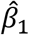 is the estimate at *t* =1. However, if the first phase *P_1_* is small, we can take 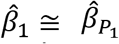 to avoid impact of extreme observation at *t* =1 (if present). As an alternative, we can take 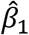 as an average of first few values of 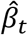. We can fit any of the following exponential functions to the estimated modifiers using least squares estimation. We have used only the first form in our study.

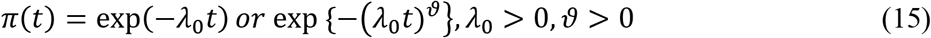

This continuous modifier function will not be phase specific and will reflect steadily increasing awareness at community-level which encourages voluntary participation in quarantine and preventive measure. The steadily decreasing modifier function can also represent the learning curve of the organizational structure associated with implementation of proposed preventive measures like quarantine, travel ban, partial lockdown and complete lockdown.

## 3 Implementation

### 3.1 Data

Since some states in India have not reported any cases and some have reported only few, we have considered populations of states with at least 10 confirmed cases reported till 20 April, 2020 for calculating total number of susceptibles. Baseline state-wise population data is obtained from the 2011 census of India (www.censusindia.gov.in). The estimated average growth rate based on the current total population of India and the total population of India in 2011 is estimated as 1.23% per annum. This rate is used to estimate current total populations of the 25 states which have been included in the calculation of total number of susceptibles. Data on the timeline of implementation of travel restrictions, isolation, lockdown, quarantine and other preventive measures by the central and state governments is compiled from various notifications issued by the Ministry of Home Affairs (MHA) and the Ministry of External Affairs (MEA) available on their official websites. Time-series data on daily prevalence of total confirmed, total recovered and total deaths is sourced from the github repository of the Centre for Systems Science and Engineering, Johns Hopkins University (https://github.com/CSSEGISandData/COVID-19).

### 3.2 Defining longitudinal phases based on containment protocols

While analyzing the effect of containment measures on rate of transmission of infection, it is important that we take into account the average incubation period. The mean incubation period of COVID-19, defined as the time from exposure to the onset of illness, is reported to be around 5 days by many researchers; refer Lauer *et al*. (2020); Chatterjee *et al*., (2020b); Yuan *et al*. (2020)) among others. This means that the impact of any intervention on the transmission rate can be expected to be visible only after 5 days, on an average. Given the fact that India has preferred focused group testing over random testing, it becomes important to address the expected lag in reporting of cases. So, for improving the analysis, cut-off dates for defining phases have been extended by 5 days to accommodate for the lag in effect induced by the incubation period. Complete lockdown in India came into effect on 25 March 2020. However, because of sudden loss of jobs and earnings of daily wagers, and the uncertainty looming over the extension of lockdown period, there were huge movements of migrant workers across India, with most of them trying to reach their homes. Overwhelming number of reports emerged about inter-state travels of large groups of people, with many even forced to travel hundreds of kilometers on foot. According to an article published in Business Standards, [Jha (2020)], on 31 March 2020 the central government reported in the Supreme Court that 500,000-600,000 migrants reached their villages on foot during the lockdown. However, as per news reports, most of the state governments, assisted by various NGOs, had come up with adequate relief shelters and food arrangements for the stranded migrant laborers by 30 March 2020. Also, affected states started compulsory quarantine facilities for people migrating from other states. These measures helped in containing any significant movement and ensuring implementation of complete lockdown. Citing these developments, we have assumed the effective date of implementation of complete lockdown as 31 March 2020. Adding incubation period of 5 days, the cut-off date for the third phase for our analysis is taken as 04 April 2020.

### 3.3 Modifier functions and hyper-parameters

Based on the phases defined in section 3.2, step-function modifier, *π(t)*, is estimated using equation (13). Negative Binomial TSIR models are chosen over Poisson TSIR models to find estimates of 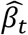 (equation (11)). Poisson models showed inflated residual deviance and proved unfit for the data. Estimated step-function is given below.

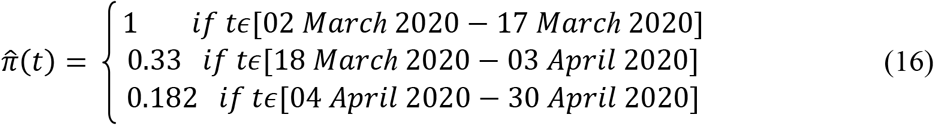

#### Hyper-parameters for Bayesian estimation

Using data on COVID-19 patients in China, Verity *et al*. (2020) have estimated mean duration from onset of symptoms to death to be 17.8 days (95% credible interval 16.9−19.2) and to hospital discharge to be 24.7 days (22.9−28.1). Mean infectious period is calculated as weighted average of these durations using observed proportions of deaths and recoveries among the total removed cases till 30 April 2020 in India as weights. The estimated mean infectious period is: 0.113 × 17.8 + 0.887 × 24.7 ≈ 24 days. Thus, the estimate for hyper-parameter for *γ* is, *γ_0_* = 1/24 = 0.042. However, since there is dearth of comprehensive studies confirming infectious period at this early stage of the epidemic, we have also performed analyses taking mean infectious period of 14 days (i.e. *γ_0_* = 1/14 = 0.0714), as reported by the World Health Organisation (WHO, 2020). So, at this juncture it is safe to assume that the reality may lie somewhere between the projections based on our two assumed cases for *γ_0_*. The value for the hyper-parameter *β* is estimated as the average of effective transmission rates over the total observed period (02 March 2020-30 March 2020). This is achieved by fitting the Negative Binomial TSIR model and using equation (12) for the entire observed period.

#### Continuous modifier function

The estimated continuous modifier function using equation (14) is given below.

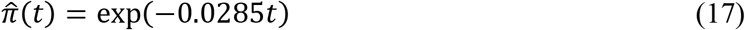

### 3.4 Forecasting assumptions

We have assumed four different scenarios for forecasting the trajectory of the COVID-19 epidemic. The four cases are summarized in Table 2. Case 1 and Case 3 are realistic scenarios based on current developments, while Case 2 and Case 4 are hypothetical scenarios strictly for the purpose of comparison.

**Table 1:** Phases of Preventive Interventions Implemented by Governments across India.

| Preventive Measures | Actual Dates of Implementation | Cut-off Dates for Defining Phases in Our Study |
| --- | --- | --- |
| No strict screening, quarantine measures, or surveillance. | 2<sup>nd</sup> March–12<sup>th</sup> March | 2<sup>nd</sup> March–17<sup>th</sup> March |
| Various social distancing measures, restrictions on public gathering, shutting down of academic institutes, international travel ban, Restriction on public transport. | 13<sup>th</sup> March–24<sup>th</sup> March | 18<sup>th</sup> March–3<sup>th</sup> April |
| Complete lockdown starts | 25<sup>th</sup> March–29<sup>th</sup> March | 4<sup>th</sup> April–30<sup>th</sup> April |
| Complete lockdown with progressive measures to tackle migrant workers' problem and containment of hotspot areas. | 30<sup>th</sup> March–19<sup>th</sup> April |  |
| Complete lockdown continues along with certain exemption for selected activities + limited movements of migrant workers within states/UT . | 20<sup>th</sup> April–3<sup>rd</sup> May |  |

**Table 2:** Cases assumed for forecasting.

| Case | Phases | Modifiers | Assumption |
| --- | --- | --- | --- |
| Case1 | P1: 2<sup>nd</sup> March–17<sup>th</sup> March | 1 | After 17 May, the effect of containment measures will remain more or less the same keeping the modifier value equal to that of the third phase, P3. |
|  | P2: 18<sup>th</sup> March–3<sup>rd</sup> April | 0.33 |  |
|  | P3: 4<sup>th</sup> April–30<sup>th</sup> April | 0.182 |  |
| Case2 | P1: 2<sup>nd</sup> March–17<sup>th</sup> March | 1 | Assuming that instead of complete lockdown only quarantine and partial lockdown measures were extended throughout after 12 March 2020. |
|  | P2: 18<sup>th</sup> March–30<sup>th</sup> April | 0.33 |  |
| Case3 | P1: 2<sup>nd</sup> March–17<sup>th</sup> March | 1 | Because of slight relaxations in green and orange zones, modifier value is assumed to increase a bit till 17 May. After that it is assumed that more economic and industrial work will start, bringing the modifier value closer to that of second phase, but will remain less than that as red zones will be strictly contained. |
|  | P2: 18<sup>th</sup> March–3<sup>rd</sup> April | 0.33 |  |
|  | P3: 4<sup>th</sup> April–3<sup>rd</sup> May | 0.182 |  |
|  | P4: 4<sup>th</sup> May–17<sup>th</sup> May | 0.2* |  |
|  | P5: 18<sup>th</sup> May and further | 0.3* |  |
| Case4 | Basic Case- No phase modifiers assumed | --- | If no containment measures were taken through the entire epidemic period. |

\* Value assumed according to the case scenario, based on the estimates of prior phases

### 3.5 Data calibration

In India, till now, testing strategy has been focused primarily on high risk individuals. However, to understand the community spread in the country, large scale random testing should be conducted among those who have no travel history [Rao *et. al*. (2020)]. As reported recently by the Indian Council of Medical Research, around 80% of the total infected (confirmed) cases in India are asymptomatic; refer www.indiatoday.in (2020). In the absence of rigorous testing, it is but natural that a large number of true cases are going undetected and hence unreported in India. This subsequently leads to concerns about the actual number of deaths due to COVID-19 also going unreported [Shaikh (2020); Biswas (2020)].

We have used a simple intuitive technique for data calibration to account for possible underreporting. We divide the observed data on total confirmed, recovered and deaths by a constant *ρ* (where 0 < *ρ* ≤ 1). Proportion of under-reporting is 1- *ρ*, i.e., *ρ* = 1 implies zero under-reporting. We have considered two levels of under-reporting, 75% (*ρ =0.25*) and 50% *(ρ=0.5)*. It is not easy to estimate the proportion of under-reporting, especially at this stage of the epidemic. However, we have based our assumptions on certain reports on scientific work in this regard; refer Jayan (2020).

### 3.6 Plotting predicted prevalence

MCMC posterior realizations on the prevalence of infected and removed are obtained from the output of *tvt.eSIR()* function of the eSIR package. Posterior mean of predicted prevalence of infecteds is plotted against time along with daily estimated prevalence of mild to moderate, severe and critical cases among the total infecteds. To predict the cases belonging to the categories mild to moderate, severe and critical, we have considered the respective proportions, 80.1%, 13.8%, and 6.1%, as reported by the World Health Organisation; refer WHO (WHO, 2020). To predict the number of deaths, we have used current proportion of deaths among the total removed cases in India, which is around 10%. Prevalence of removed is plotted against time along with estimated number of cases for the events recovered and death. Plots for case 3 of step-function modifier and for exponential modifier at two different values of *γ* are presented in Graphs 1-5.

**Graph 1:**
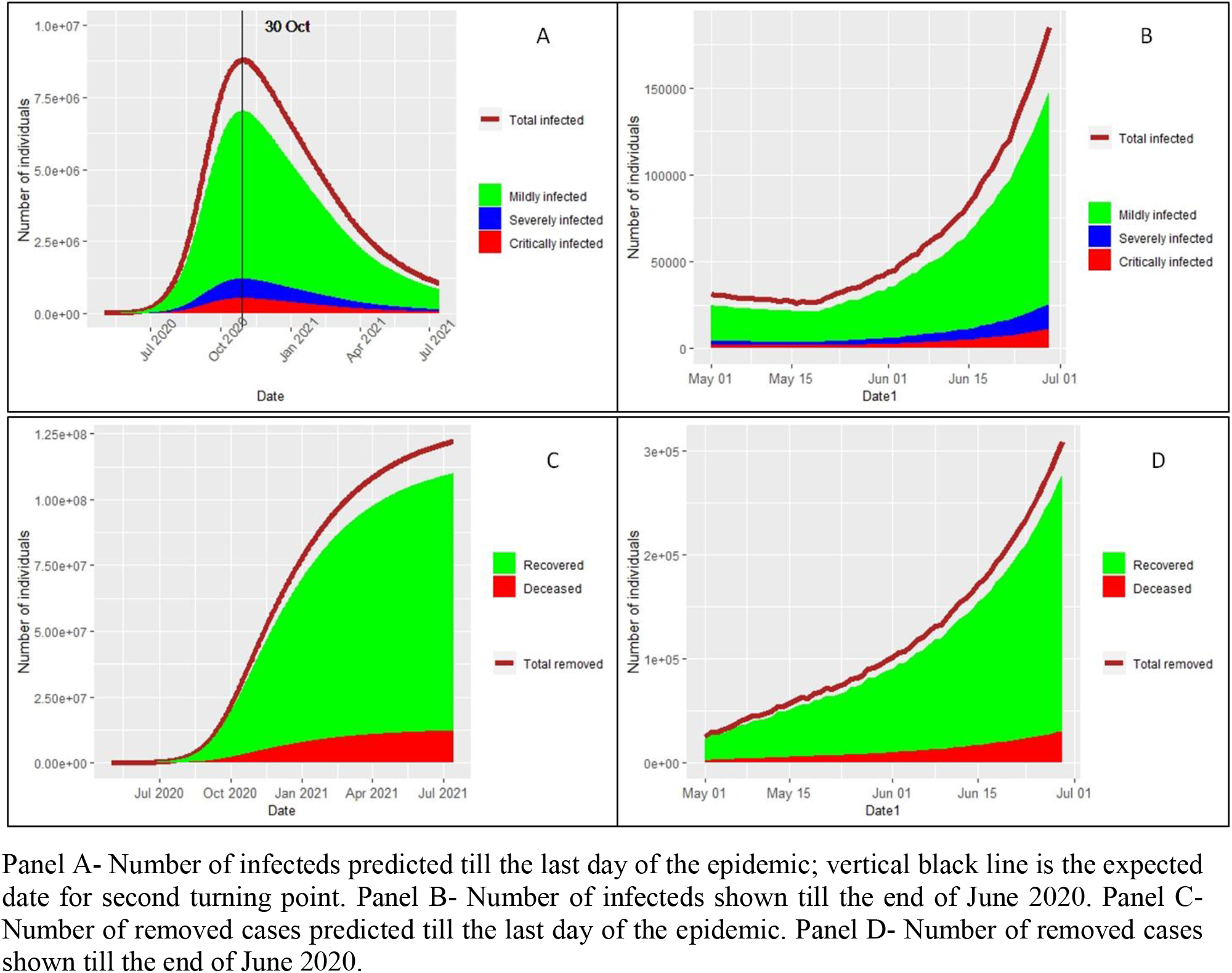
Predictions from Case 3 of step-function modifier- (at 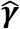 = 0.0714)

**Graph 2:**
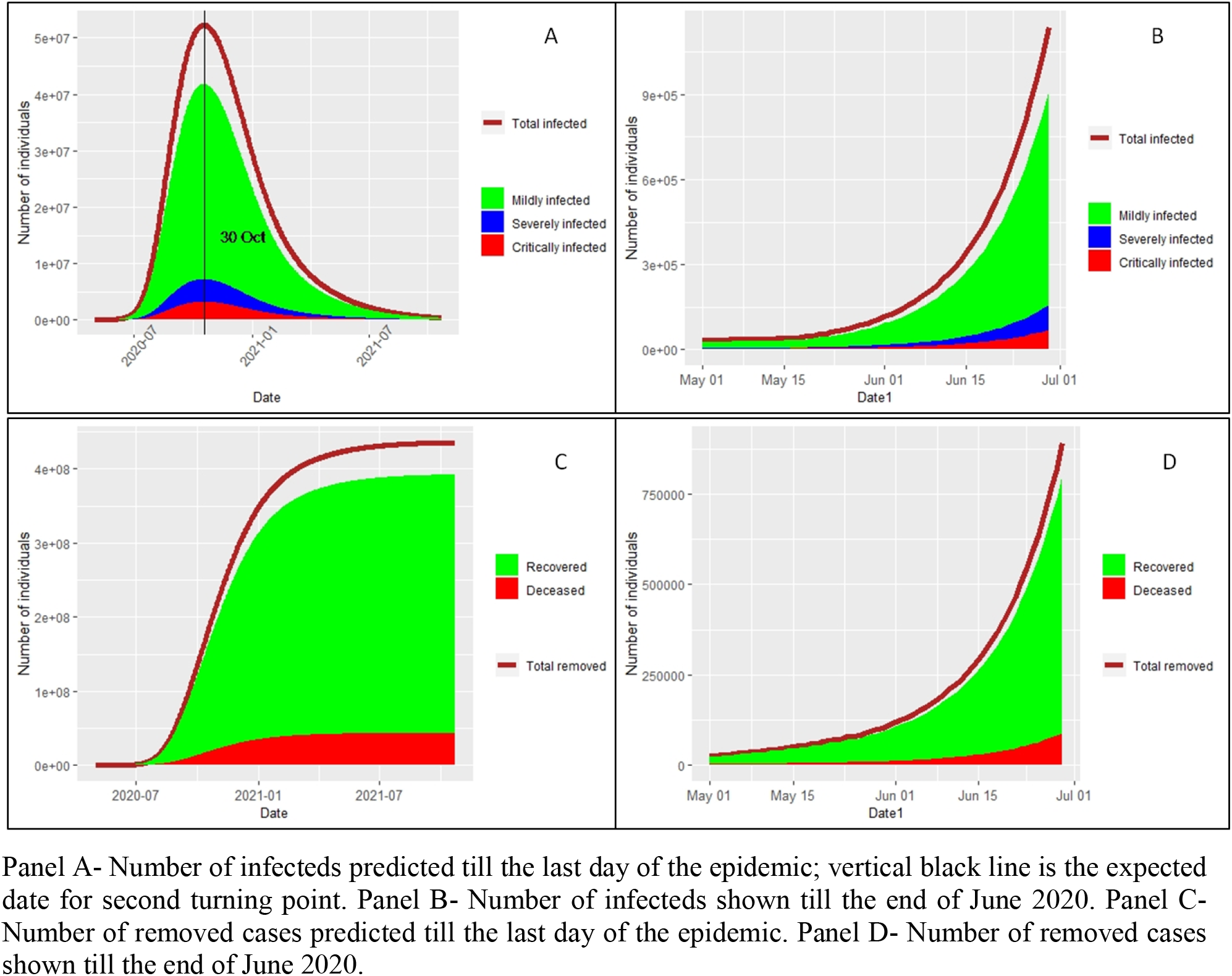
Predictions from Case 3 of step-function modifier- (at 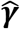 = 0.042)

**Graph 3:**
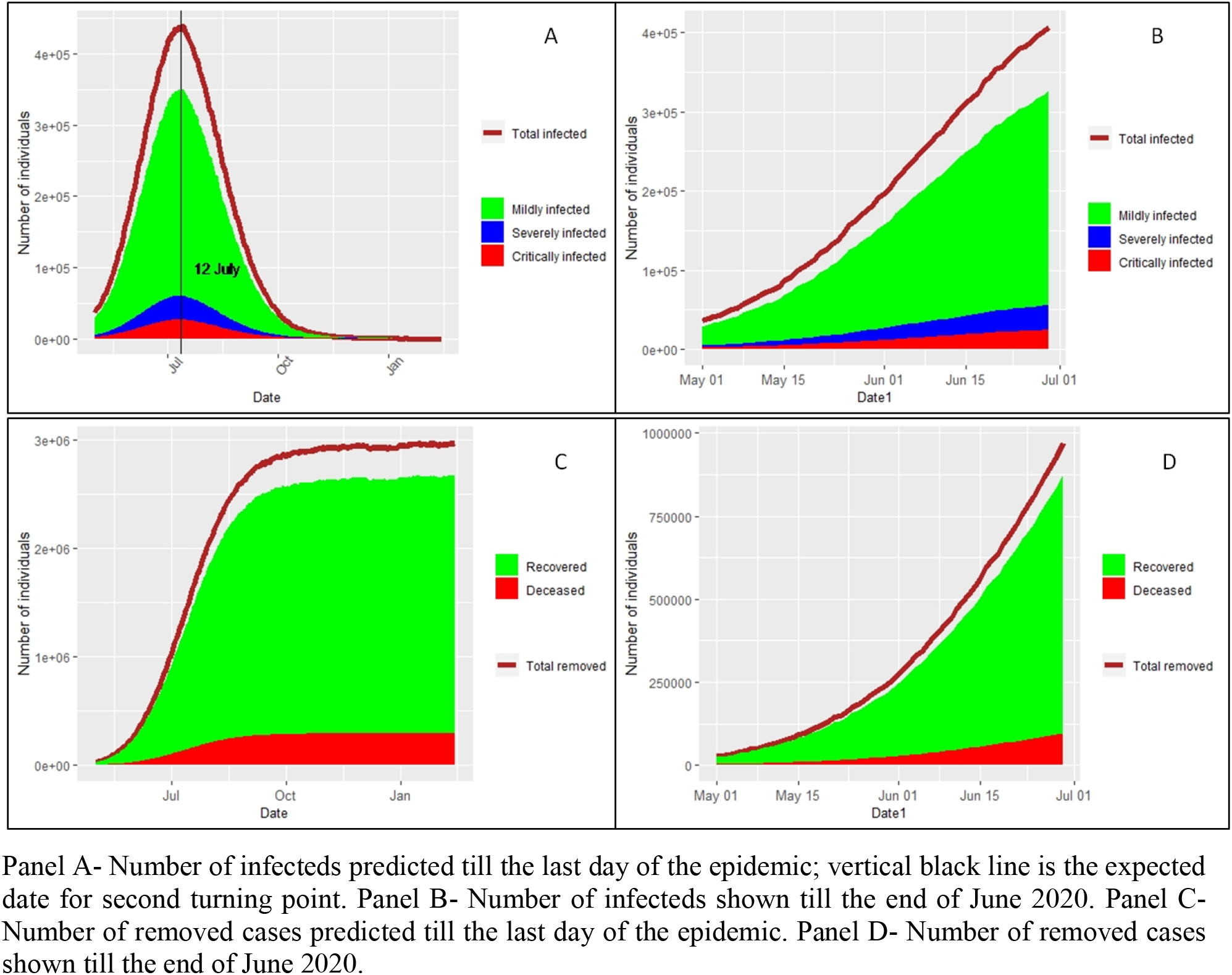
Predictions from exponential modifier function- (at 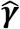 = 0.0714)

**Graph 4:**
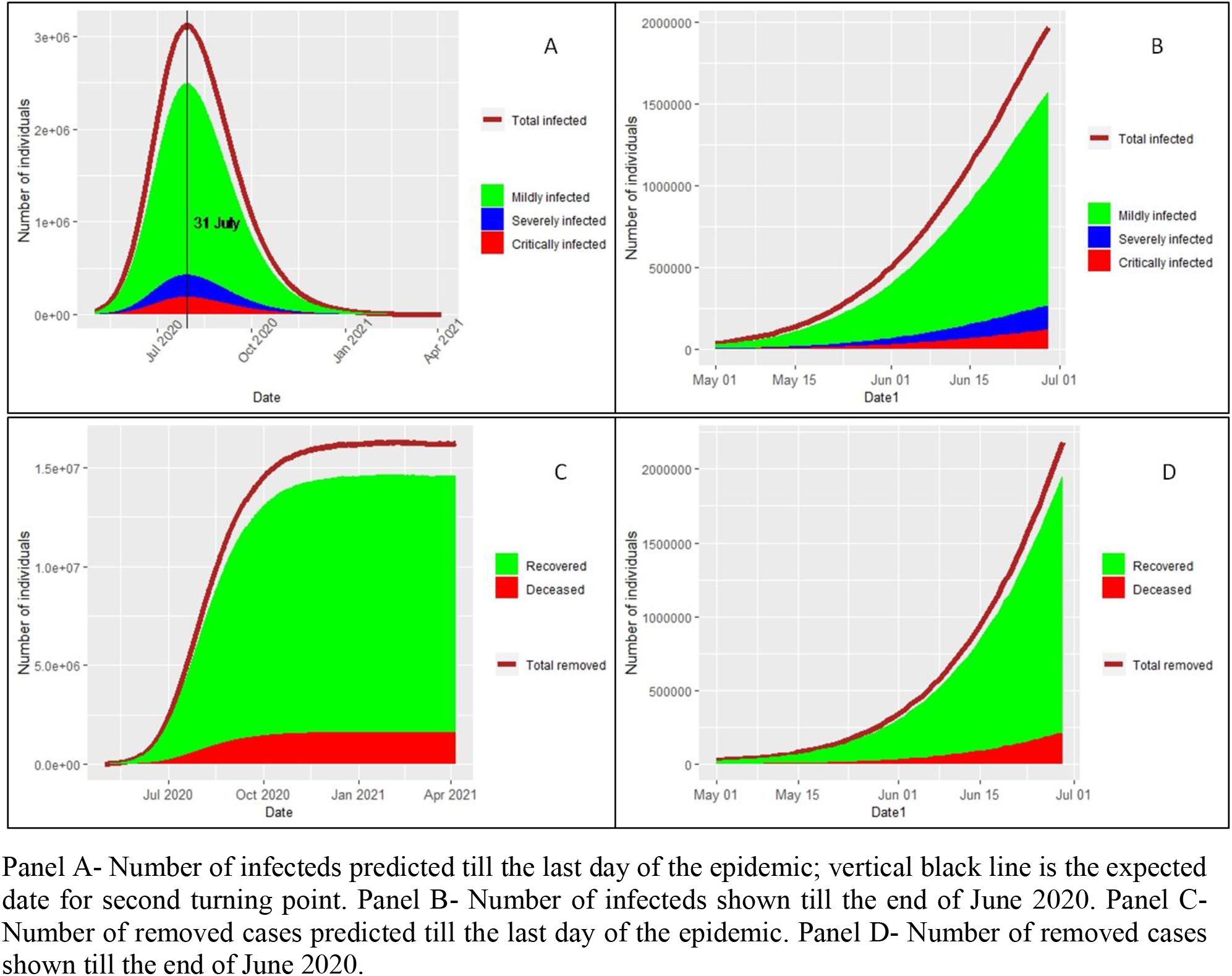
Predictions from exponential modifier function- (at 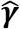 = 0.042)

**Graph 5:**
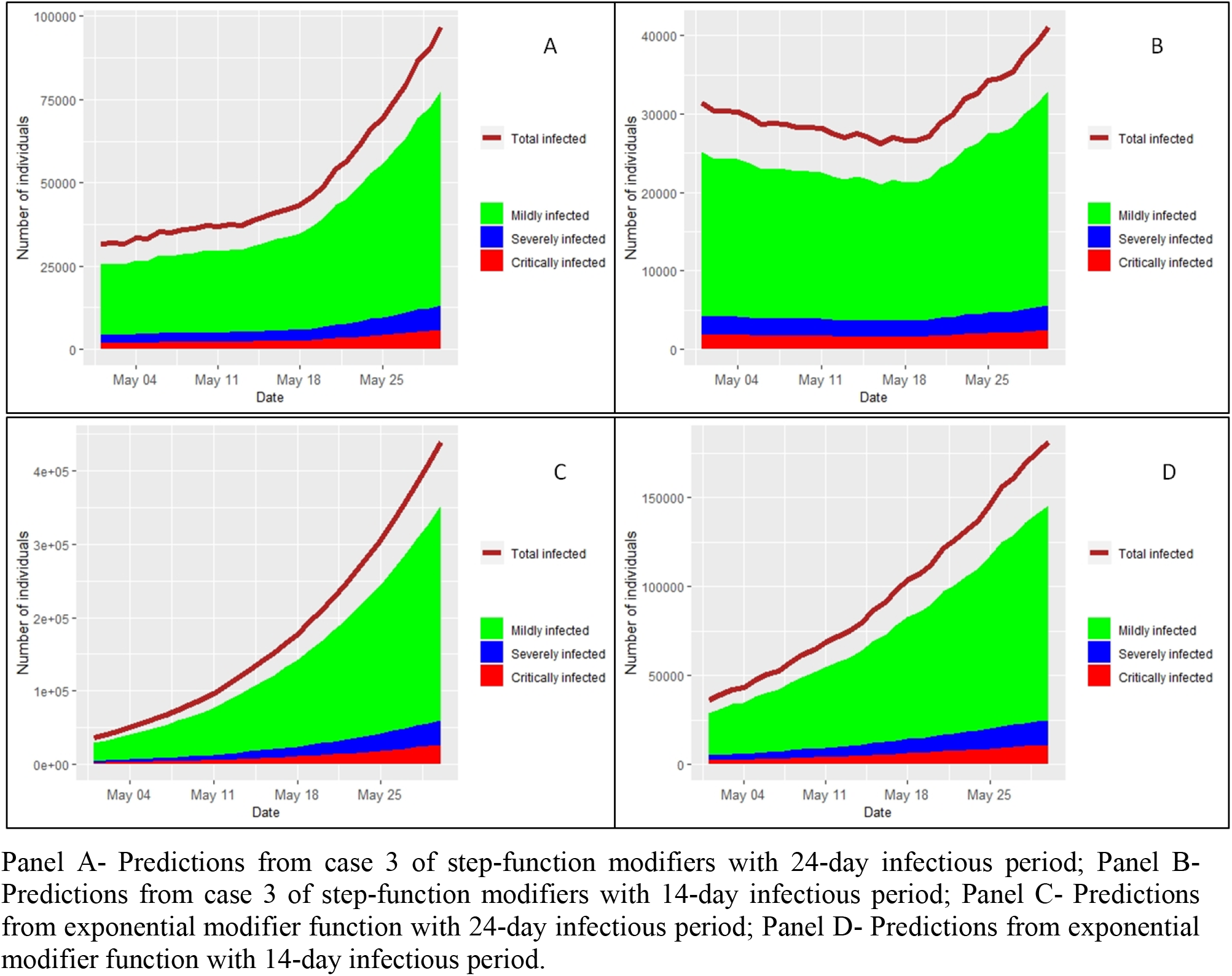
Predictions of number of infecteds for May 2020 from case 3 of step-function modifiers and exponential modifiers

## 4 Results and Discussion

Estimated values of time-dependent transmission rate adjusted for modifier, 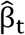 (for phase 1, 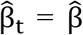), rate of removal, 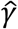, and reproduction number *R_0_*, along with their 95% credible intervals based on posterior realizations are reported for all models and all cases discussed in the implementation section. Expected total incidence (as % of total population), and forecasted dates for two crucial turning points of the epidemic are also reported for each case. The first turning point signifies the time at which the rate of increase in the number of infecteds starts decreasing (deceleration). The second crucial turning point is the peak time of the infected curve beyond which the prevalence of infecteds starts decreasing. Table 3 and Table 4 present results for all four cases of step-function modifier based eSIR, for the initial estimate of infectious period as 24 days and 14 days respectively. Results from the exponential modifier function based eSIR, for both observed and calibrated data, are presented in Table 5. Table 6 contains results for calibrated data using case 3 of step-function modifier in the eSIR model.

**Table 3:**
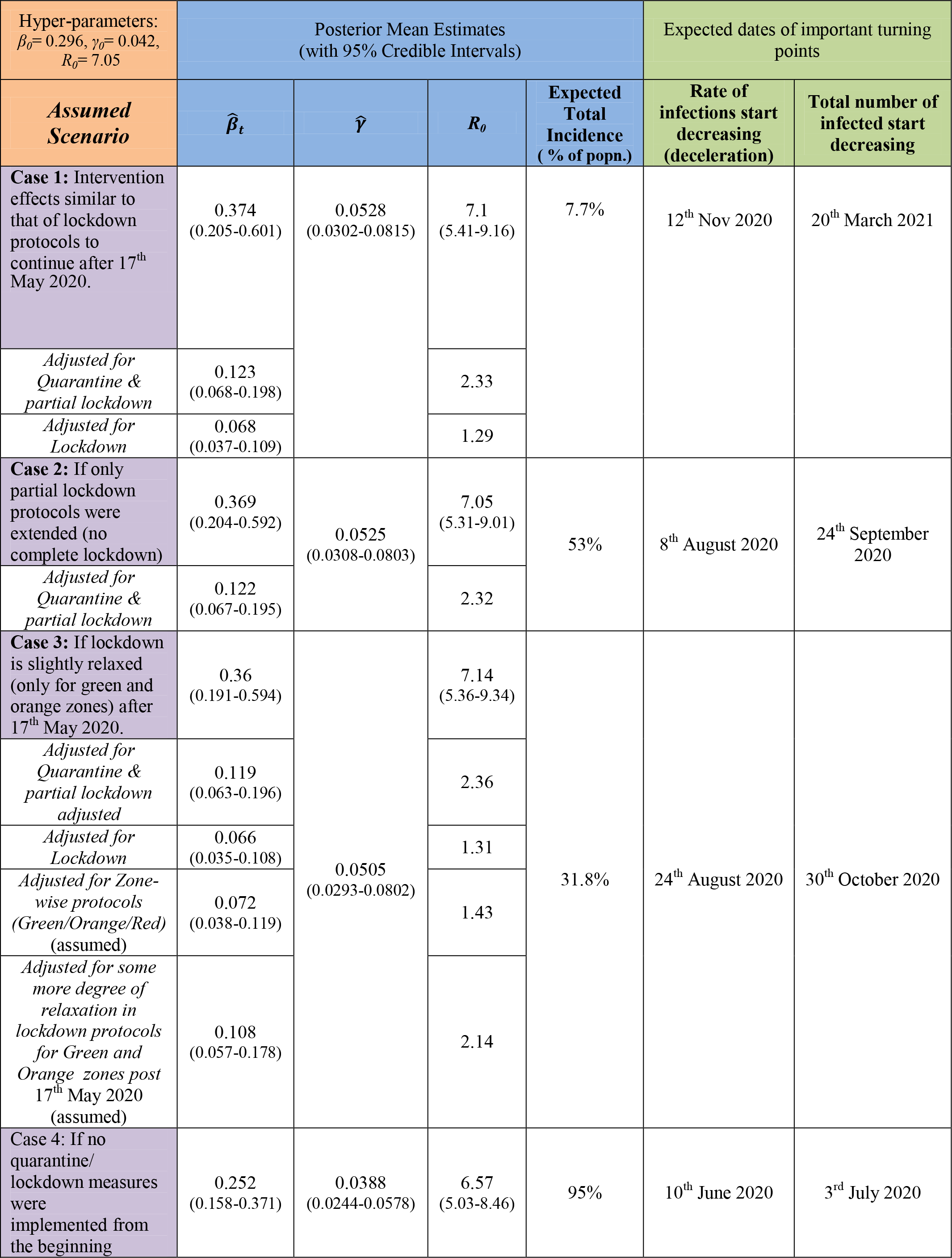
Estimated/ predicted results using step-function modifiers (Infectious period = 24 days)

**Table 4:**
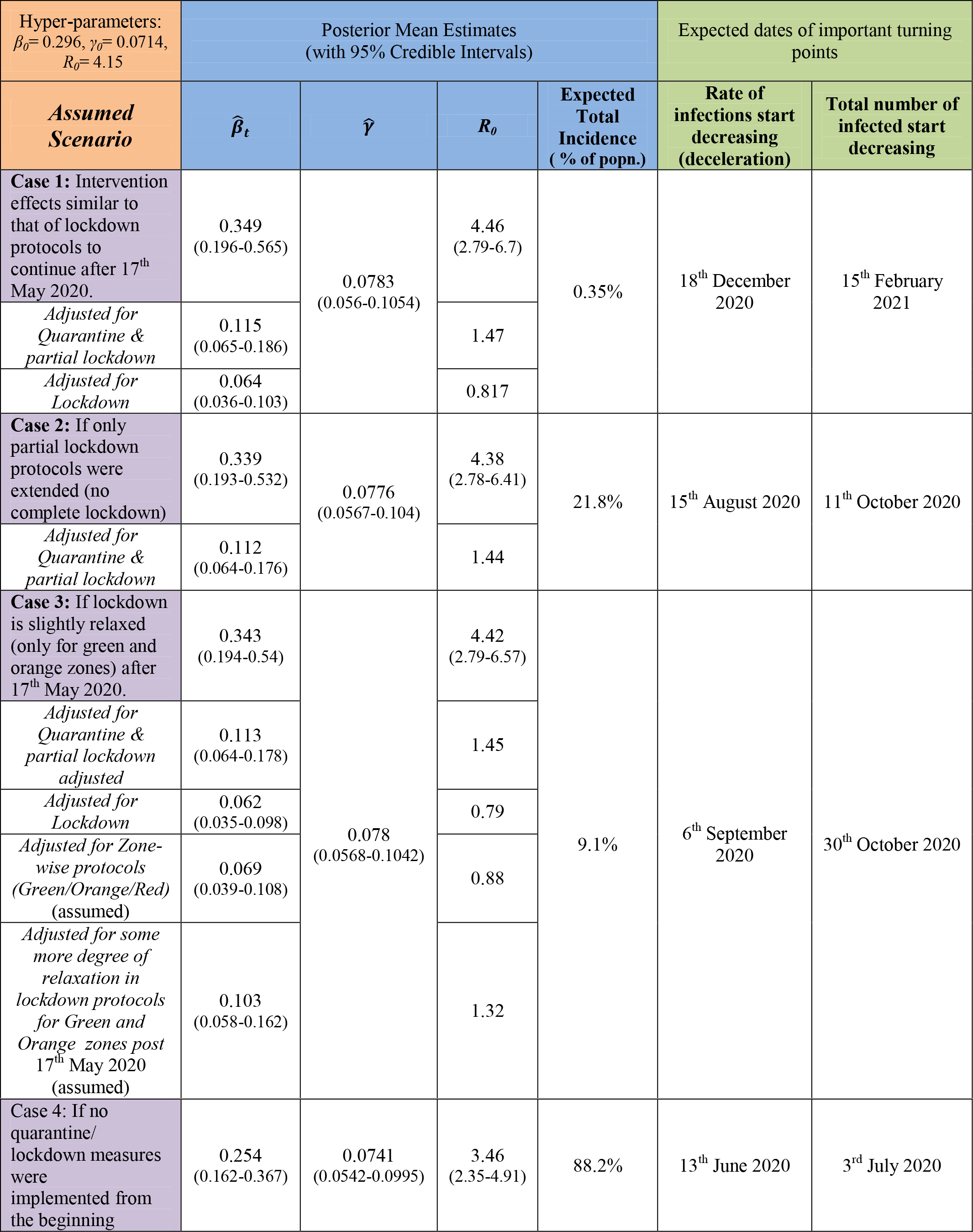
Estimated/ predicted results using step-function modifiers (Infectious period = 14 days)

**Table 5:** Estimated/ predicted results using exponential modifier function.

| $\lambda_0 = 0.0131$ | Posterior Mean Estimates<br>(with 95% Credible Intervals) | | | | Expected dates of important turning points | |
| --- | --- | --- | --- | --- | --- | --- |
| Case | $\hat{\beta}$ | $\hat{\gamma}$ | $R_0$ | Expected Total Incidence<br>( % of popn.) | Rate of infections start decreasing<br>(deceleration) | Total number of infected start decreasing |
| Without data calibration (assuming scale of under-reporting is not significant) |  |  |  |  |  |  |
| Hyper-parameters:<br>$\beta_0 = 0.296, \gamma_0 = 0.042,$<br>$R_0 = 7.05$ | 0.329<br>(0.19-0.509) | 0.0473<br>(0.0287-0.0767) | 7.01<br>(5.24-8.94) | 1.2% | 26<sup>th</sup> June 2020 | 31<sup>st</sup> July 2020 |
| Hyper-parameters:<br>$\beta_0 = 0.296, \gamma_0 = 0.0714,$<br>$R_0 = 4.15$ | 0.326<br>(0.19-0.504) | 0.0788<br>(0.0566-0.1046) | 4.16<br>(2.65-6.07) | 0.22% | 8<sup>th</sup> June 2020 | 12<sup>th</sup> July 2020 |
| With data calibration for under-reporting (assuming 75% under-reporting) |  |  |  |  |  |  |
| Hyper-parameters:<br>$\beta_0 = 0.296, \gamma_0 = 0.042,$<br>$R_0 = 7.05$ | 0.312<br>(0.196-0.458) | 0.0466<br>(0.0293-0.0694) | 6.76<br>(5.2-8.71) | 2% | 22<sup>nd</sup> June 2020 | 29<sup>th</sup> July 2020 |
| Hyper-parameters:<br>$\beta_0 = 0.296, \gamma_0 = 0.0714,$<br>$R_0 = 4.15$ | 0.307<br>(0.187-0.443) | 0.0751<br>(0.0564-0.0981) | 4.11<br>(2.78-5.73) | 0.38% | 30<sup>th</sup> May 2020 | 2<sup>nd</sup> July 2020 |
| With data calibration for under-reporting (assuming 50% under-reporting) |  |  |  |  |  |  |
| Hyper-parameters:<br>$\beta_0 = 0.296, \gamma_0 = 0.042,$<br>$R_0 = 7.05$ | 0.326<br>(0.192-0.491) | 0.0481<br>(0.0292-0.0727) | 6.83<br>(5.22-8.86) | 1.65% | 24<sup>th</sup> June 2020 | 30<sup>th</sup> July 2020 |
| Hyper-parameters:<br>$\beta_0 = 0.296, \gamma_0 = 0.0714,$<br>$R_0 = 4.15$ | 0.322<br>(0.192-0.475) | 0.0774<br>(0.0566-0.1050) | 4.18<br>(2.67-5.9) | 0.28% | 4<sup>th</sup> June 2020 | 8<sup>th</sup> July 2020 |

**Table 6:**
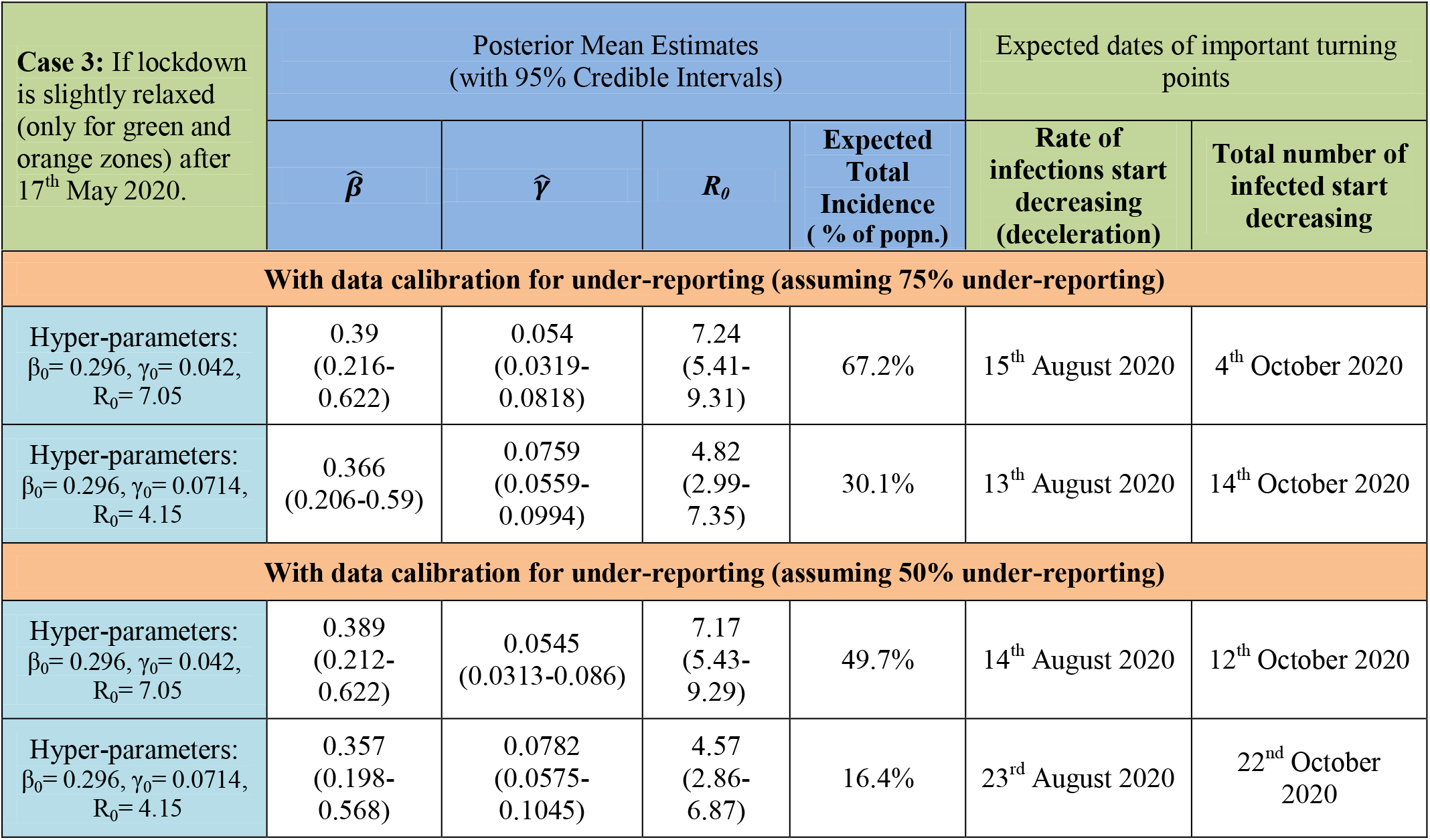
Estimated/ predicted results using step-function modifiers and calibrated data.

| Case 3: If lockdown is slightly relaxed (only for green and orange zones) after 17<sup>th</sup> May 2020. | Posterior Mean Estimates<br>(with 95% Credible Intervals) |  |  |  | Expected dates of important turning points |  |
| --- | --- | --- | --- | --- | --- | --- |
| | $\hat{\beta}$ | $\hat{\gamma}$ | $R_0$ | Expected Total Incidence<br>( % of popn.) | Rate of infections start decreasing<br>(deceleration) | Total number of infected start decreasing |

| With data calibration for under-reporting (assuming 75% under-reporting) |  |  |  |  |  |  |
| --- | --- | --- | --- | --- | --- | --- |
| Hyper-parameters:<br>$\beta_0 = 0.296$, $\gamma_0 = 0.042$,<br>$R_0 = 7.05$ | 0.39<br>(0.216-0.622) | 0.054<br>(0.0319-0.0818) | 7.24<br>(5.41-9.31) | 67.2% | 15<sup>th</sup> August 2020 | 4<sup>th</sup> October 2020 |
| Hyper-parameters:<br>$\beta_0 = 0.296$, $\gamma_0 = 0.0714$,<br>$R_0 = 4.15$ | 0.366<br>(0.206-0.59) | 0.0759<br>(0.0559-0.0994) | 4.82<br>(2.99-7.35) | 30.1% | 13<sup>th</sup> August 2020 | 14<sup>th</sup> October 2020 |
| With data calibration for under-reporting (assuming 50% under-reporting) |  |  |  |  |  |  |
| Hyper-parameters:<br>$\beta_0 = 0.296$, $\gamma_0 = 0.042$,<br>$R_0 = 7.05$ | 0.389<br>(0.212-0.622) | 0.0545<br>(0.0313-0.086) | 7.17<br>(5.43-9.29) | 49.7% | 14<sup>th</sup> August 2020 | 12<sup>th</sup> October 2020 |
| Hyper-parameters:<br>$\beta_0 = 0.296$, $\gamma_0 = 0.0714$,<br>$R_0 = 4.15$ | 0.357<br>(0.198-0.568) | 0.0782<br>(0.0575-0.1045) | 4.57<br>(2.86-6.87) | 16.4% | 23<sup>rd</sup> August 2020 | 22<sup>nd</sup> October 2020 |

At *γ_0_* = 0.042 (24-day infectious period), estimated values of the production rate R_0_ (unadjusted) consistently stays around 7 in all cases, and at *γ_0_* = 0.0714 (14-day infectious period) its estimates cluster around 4 for all cases. Consistency of the estimate of *R_0_* (unadjusted) and *β* (base value unadjusted for modifiers) under different hypothesized situations suggests that our estimates of modifiers are able to explain the changes in the transmission rate in their respective phases. Estimates of *R_0_* are comparatively very small in the containment phases, with that for the complete lockdown being the minimum. For example, citing results of case 1 from Table 3, under the assumption of 24-day infectious period, *R_0_* is estimated to be 1.29 for the lockdown period, and around 2.33 for the quarantine/ partial lockdown phase, as opposed to 7.1 in the no-intervention phase. Similar results are obtained for the case 3, at both levels of *γ_0_. R_0_* values estimated from exponential modifier function based approach are slightly on the lower side as compared to those obtained from the step-function approach. This is expected as the use of exponential modifier function results in continuous decline in the transmission rates through time.

Estimate of (mean) total incidence is very sensitive to the choice of infectious period. Even under the assumption of complete lockdown till the end of epidemic, the estimated total incidence jumps from 0.35% to 7.7% of the population as we increase the infectious period from 14 days to 24 days. Similar jumps are seen in all cases. Unfortunately, as discussed in section 3.3, the existing reports on COVID-19 at this early stage of the epidemic are not conclusive about the duration of infectious period. In addition, current recovery/ death trends of different countries indicate that recovery rates and death rates can vary significantly between different regions.

Although we obtained most optimistic results under the assumption of complete lockdown like situation throughout the course of the pandemic, it is not practical to believe that our economy can sustain such a measure for such a long duration. Among the situations assumed for prediction using step-function modifiers, future assumptions for case 3 are practically most achievable. Also, daily predictions of number of infecteds for the month of May based on case 3 of step-function modifiers, with 24-day infectious period, are closest to the actual reported data as compared to those of any other scenario considered in this study; refer Graph 5. To restrict the COVID-19 spread within the limits predicted by case 3 results, we have to ensure that the post complete lockdown period should not let R_0_ to go beyond 1.32 (14-day infectious period) or beyond 2.14 (24-day infectious period). If we are able to do that, depending on the actual recovery time of COVID-19 patients in India we can expect around 9.1% to 31.8% of the total population to get infected with SARS-CoV-2 by the time the epidemic ends. Assuming 75% under-reporting of infected and recovered/ deceased cases, the range of expected total incidence becomes 30.1% - 67.2%, and for 50% under-reporting of cases, it is estimated as 16.4% - 49.7%. The rate of infection is expected to start decreasing around the end of August or start of September 2020, and the total number of active cases is expected to start declining towards the end of October 2020. If there is under-reporting of cases, these dates of turning points are expected to shift earlier by around a week.

It is also worth mentioning that if the lockdown measures had not been implemented and only quarantine and partial lockdown were continued (case 2), we would be expecting around 22% to 53% of the population to be infected by the end of the epidemic. And if there was no containment measure in place since start (case 4), 88% to 95% of the population would have contracted the infection till the time epidemic lasted.

Since the exponential modifier function assumes a continuous decline in the effective transmission rate, it may overlook some important real life factors while predicting the course of the epidemic, and hence it may result in underestimation of the overall impact of the epidemic. As expected, the results obtained using this approach is closer to those of case1 where complete lockdown is assumed beyond 17 May 2020. Use of exponential modifier function may not be the best way to describe the effects of sudden drastic measures like complete lockdown, travel ban *etc*.

Even in the best case scenario as depicted by the results obtained using the exponential modifier function, the total incidence is predicted to be up to 1.2% (without data calibration) and up to 2% (with data calibration). That is, around 16 million to 27 million people are expected to end up getting infected with SARS-CoV-2 by the time the epidemic ends. According to the predictions from the exponential modifier approach, when the prevalence of infected reaches its peak around mid to end of July 2020, there will be around 60,000 (14-day recovery period) to 500,000 (24-day recovery period) severe cases who will need hospitalization, at once (Graph 3 and Graph 4). The picture becomes even more unsettling when we study the graphs of the case 3 predictions (Graph 1 and Graph 2). These figures range between 1 million to 8 million for the two cases, and the peak time is expected to be around the end of October 2020.

## 6 Conclusion

Substantial reduction in the reproduction rate *R_0_* during the partial lockdown and complete lockdown phases corroborates the effectiveness of these interventions in containing the spread of SARS-CoV-2 infection. Assuming an average recovery (or infectious) period of 14 days, *R_0_* is estimated to have reached below 1 in the complete lockdown phase. However, assuming a 24-day recovery period, the estimate of *R_0_* remained above 1 even during the complete lockdown. In case 3 we have considered existing situation of containment measures till 17 May 2020, and at 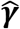 (24-day infectious period) the daily predictions for May 2020 are much closer to the actual reported values (as observed till 11 May 2020) as compared to those at 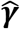 (14-day infectious period). However, more clinical reports based on wider patient level data are imperative towards finding reliable estimates of recovery time for COVID-19 patients.

The fact that instead of using pre-specified modifier for each phase, we have estimated phase-specific modifiers from the observed data improves our chances of obtaining more reliable estimates of transmission rate and *R_0_* as compared to other recent studies on India, like Ray *et al*. (2020). Use of lower than true values of modifiers may lead to over-estimation of transmission rate and vice-versa. Also, our procedure of defining cut-off dates for different phases of containment measures assimilates the effects of incubation period and initial lapses in the implementation of the lockdown.

Even under the most optimistic scenarios, the time for flattening of curve is still quite far. The number of infected cases is expected to increase at even a higher rate at the moment and by the time the peak is expected, we will need an extensive amount of medical and infrastructural preparedness. Quoting the results based on the assumptions of case 3, which we have repeatedly deemed as the most realistic case, and whose predictions for the first week of May 2020 are closest to the observed values among all cases, we need to be prepared with enough medical infrastructure and equipments to be able to handle between 1 million to 8 million severe cases around the end of October 2020.

### Limitations

We have considered same estimates of phase-specific modifier for the entire country assuming that the lockdown and other containment protocols have been homogeneously implemented across India. However, because of significant differences in various socio-economic, demographic, cultural, and administrative level factors, actual transmission rates are bound to differ from region to region. Hence, the estimated parameters in our study are only valid for overall predictions of cases in India, on an average, and may fail to trace the dynamics of the epidemic in sub-regions, say districts or states.

## Data Availability

github repository of the Centre for Systems Science and Engineering, Johns Hopkins University

https://github.com/CSSEGISandData/COVID-19

## Appendix-A Detailed time-line of containment measures implemented in India

18.01.2020: Thermal screening of all passengers coming from China and Hong kong started at three international airports.
30.01.2020: First Covid-19 case reported in India. Travel history from Wuhan, China
04.03.2020: By this date thermal screening was initiated in a progressive manner (depending on spread of the disease to other countries) for all international passengers at all port of entry (land, sea and air-ports) through various travel advisories.
13.03.2020 - 22.03.2020: During this period various state governments brought out notices for restricting social contacts - ban on public gatherings of any kind; shutting down of academic institutes; restrictions on public transportation; screening of interstate passengers at airports; and complete lockdown of some states from 23.03.2020 till 31.03.2020.
15.03.2020 – 21.03.20202: Min. 14-days quarantine made mandatory for all incoming travelers from covid-19 infected countries (in progressive manner). Also suspension of all Visa till 15/04/2020
22.03.2020: Ban on all incoming international flights, except those already on transit. Suspension of all mass transportation services, like metro, rail, domestic air, till 31/03/2020, except those that started their journey before 22.03.2020.
25.03.2020: Complete lockdown till 14/04/2020. However, large-scale movements of migrant workers across various states started from 26/03/2020.
29.03.2020: Order issued to all state governments on this date to stop the migrants’ movements and setting up of relief camps for those already in transit.

From the above box, we can take:

30/03/2020: As effective date of starting of measures to stop migrant movements across various states during complete lockdown till 14/04/2020.
15/04/2020: Complete lockdown extended till 03/05/2020 with new containment measures for hotspot areas.
20/04/2020: Complete lockdown till 03/05/2020 with new containment measures for hotspot areas + certain exemption for selected activities + limited movements of migrant workers with states/UT.

